# Roles of beta synchronisation for motor skill acquisition change after stroke

**DOI:** 10.1101/2025.10.01.25337084

**Authors:** Lena S. Timmsen, Benjamin Haverland, Silke Wolf, Charlotte J. Stagg, Jan Feldheim, The Vinh Luu, Robert Schulz, Till R. Schneider, Fanny Quandt, Bettina C. Schwab

**Author notes:** These authors contributed equally to this work (Fanny Quandt, Bettina C. Schwab). Corresponding authors: Bettina C. Schwab, University of Twente, Drienerlolaan 5, 7522 NB Enschede, The Netherlands,; Fanny Quandt, University Medical Center Hamburg-Eppendorf, Martinistraße 52, 20246 Hamburg, Germany.

## Abstract

Beta event-related synchronisation (ERS) following movement has been associated with motor skill acquisition in healthy individuals, yet its role in stroke recovery remains unclear. Given the prevalence of motor impairments after stroke, understanding how beta ERS relates to motor skill acquisition in this population is of significant clinical relevance, especially in view of emerging opportunities for neuromodulation.

In this study, we investigated whole-brain beta ERS during a feedback-guided motor skill acquisition task using magnetoencephalography (MEG) in 14 well-recovered stroke survivors in the chronic phase and 15 age-eligible healthy control participants. Motor ability was assessed with standardized clinical scales, and structural brain metrics were derived from magnetic resonance imaging. MEG data were projected into source space to enable comprehensive cluster-based analyses across the cortex.

While stroke survivors exhibited significantly lower overall task performance, their capacity for motor skill acquisition did not significantly differ from that of control participants. In healthy participants, motor skill acquisition was strongly and positively associated with beta ERS in a cluster encompassing bilateral sensorimotor areas, which was absent in stroke survivors. Instead, stroke survivors showed a trend towards a negative association of motor skill acquisition with beta ERS. An exploratory analysis revealed that among various clinical and structural measures, only the Box and Block Test, a measure of gross manual dexterity, significantly moderated the association between beta ERS and motor skill acquisition.

These findings suggest that although stroke survivors may retain the ability to acquire motor skills, the underlying neural mechanisms can be altered. In healthy adults with high manual dexterity, beta ERS appears to support short-term motor skill acquisition, whereas at lower dexterity levels, alternative mechanisms may compensate to sustain skill acquisition.

**Abbreviated Summary:** Timmsen et al. use functional brain imaging to show that although stroke survivors retain the ability to obtain new motor skills, the brain activity supporting this process differs from that of healthy individuals, suggesting that alternative neural mechanisms may compensate after stroke.

**Graphical Abstract:** 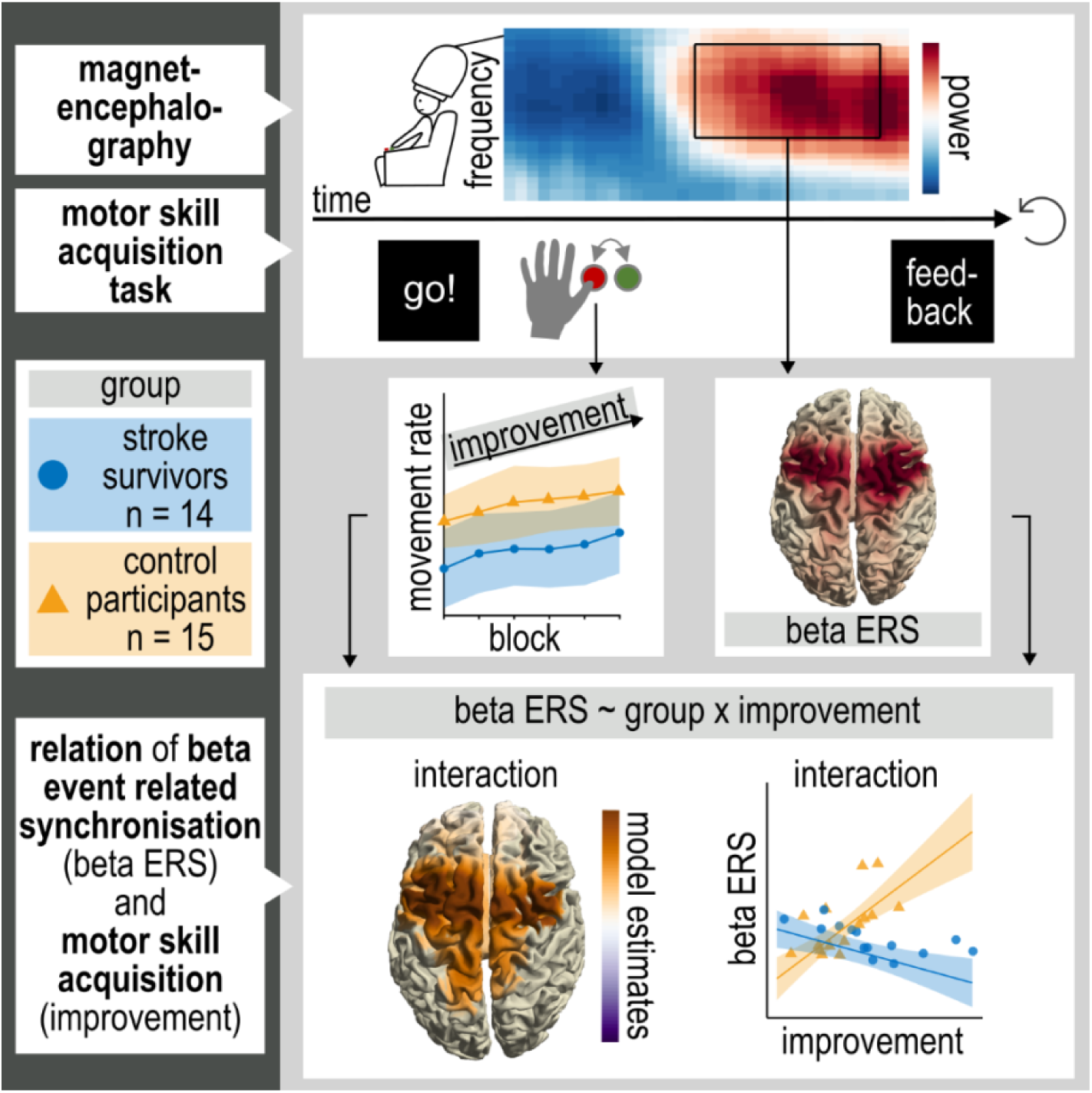

## Introduction

Neural oscillations are considered critical for coordinating neuronal activity across spatially distributed brain regions, thereby enabling the integration of information required for complex motor behaviours.^1^ Specifically, movement-related beta activity (13–30 Hz) may reflect intracortical inhibition and play a key role in motor preparation and execution.^2,3^ They are characterized by a suppression of beta power before and during movement, referred to as event-related desynchronisation (beta event-related desynchronisation/beta ERD), followed by a post-movement rebound (beta event-related synchronisation/beta ERS). Beta ERD is typically associated with motor preparation or active movement, indicating cortical activation and reduced synchrony among neuronal populations.^2,4^ In contrast, beta ERS is often linked to motor inhibition or sensory processing, reflecting cortical idling or the re-establishment of resting-state synchrony.^5,6^ Beyond reflecting the current motor status, beta activity also plays a role in motor learning and skill acquisition,^7–11^ and beta ERS has been shown to relate to movement error.^12–14^ After a stroke, movement-related beta activity can be altered. Beta ERD and ERS are reduced,^15–17^ and the extent of reduction has been associated with greater motor impairment.^15,17^ Depending on the stroke cohort, alterations in ERD levels over time have been associated with the recovery process.^18^ Also, the beta rebound increased with recovery over time,^16^ and its reduction persists even in individuals after stroke with only minor impairment.^19,20^ While stroke survivors generally keep their ability to acquire new motor skills,^21,22^ the relationship of beta ERS and motor skill acquisition after stroke remains less clear. Existing research on this topic includes a single study^23^, which demonstrated that well-recovered stroke survivors exhibited diminished modulation of beta activity in response to training. In that study^23^ the ERS after training further contributed to the prediction of motor performance 24 h after training. These findings suggest that movement-related beta activity may not only be a marker of motor function but could also be indicators of motor skill acquisition capacity and neural plasticity following stroke.

Understanding the mechanisms of motor control and skill acquisition after partial loss of function after stroke could play an essential role in designing new approaches for therapy. If causal mechanisms of stroke recovery would be known, they would allow for the individualized design of neuromodulation strategies for rehabilitation. Specifically, brain stimulation methods like transcranial alternating current stimulation (tACS) or transcranial magnetic stimulation (TMS) may be tailored to re-adjust pathological dynamics and help to regain motor function as soon as possible.^24^

In this study, we focused on how short-time motor skill acquisition, measured within a single day, relates to beta activity at the whole-brain level. To this end, we applied a motor skill acquisition task based on thumb abduction, which was successfully modulated by tACS in healthy participants and stroke survivors.^25,26^ Unlike many prior studies that restrict analyses to predefined regions of interest, we used a data-driven approach combining high-resolution MEG source reconstruction combined with motor behaviour. This allowed us to identify spatial clusters where beta activity was associated with motor skill acquisition, without the bias of a priori region selection. Given the diverse roles of beta ERS in both general motor control and skill learning, we hypothesized that these roles vary across cortical regions and may be altered following stroke. To test this, we re-analysed previously acquired behavioural, MEG, and magnetic resonance imaging (MRI) data, along with clinical scores, from stroke survivors in the chronic phase of recovery and age-eligible control participants.^27^

## Materials and methods

### Participants and setting

As previously described^27^, we recruited 16 stroke survivors in the chronic phase and 18 healthy control participants. The control participants were matched for the mean and standard deviation of the stroke survivors’ age distribution. The stroke survivors had experienced a first-ever clinical stroke at least six months prior to participation, exhibited an upper extremity deficit lasting a minimum of 24 hours, and presented with a structural lesion visible on clinical MRI or computed tomography. All participants were right-handed, as determined by the Edinburgh Handedness Inventory. Individuals taking psychotropic medication were excluded. Three control participants were excluded retrospectively due to non-compliance with inclusion criteria, specifically pre-existing neurological conditions and left-handedness. Additionally, two stroke survivors were excluded due to significant MEG artifacts associated with metal implants.

The data set comprised MEG measurements collected during motor task execution, behavioural data, structural MRI scans, and clinical scores. Stroke survivors and control participants both received standardized clinical testing with the Fugl-Meyer Assessment of the Upper Extremity (UEFM), Action Research Arm Test (ARAT), Nine Hole Peg Test (NHPT), Box and Block Test (BBT), whole hand grip force, key grip force, National Institutes of Health Stroke Scale (NIHSS), modified Rankin Scale (mRS) and Mini Mental State Examination (MMSE).

The study was conducted according to the Declaration of Helsinki, except for preregistration. Participants gave written informed consent. The local ethics committee of the Medical Association of Hamburg approved the study (2021-10410-BO-ff).

### Motor Skill Acquisition Task

Participants were comfortably seated in the MEG system, facing a screen, with their performing arm secured in a splint. A two-button response box (Current Designs, Philadelphia, PA) was positioned so that the participant’s thumb could reach both buttons. In response to a “ready – steady – go” cue, participants were instructed to press the two buttons alternately for a total of four button presses using their thumb, while keeping their arm relaxed (Fig. 2A).

Participants completed 20 familiarisation trials before they were instructed in a standardized way to perform the task as fast as possible. Each participant then completed six blocks, each consisting of 40 trials. After each trial, participants received feedback on their absolute performance as well as on their improvement relative to both the current and the previous blocks. At the end of each block, additional feedback was provided in the form of a bar graph indicating average performance for all completed blocks. If a participant pressed a button before the go-cue, took longer than a second to start the first button press, or pressed the buttons in an incorrect order, an error message was shown. Stroke survivors performed the task with the hand contralateral to their lesioned hemisphere, and the performing hand of control participants was selected to match that of the stroke survivors.

### Data acquisition

#### MEG and MRI data acquisition

We recorded MEG data with a CTF-MEG system (CTF Systems, Coquitlam, Canada, 275 axial gradiometers, sampling rate 1200 Hz), including a continuous measurement of the head position.^28^ Participants were instructed to return to their initial head position after every block. We furthermore recorded the electrooculogram (horizontal and vertical) and electrocardiogram via bipolar channels, and time points of button presses and visual stimuli via LPT triggers.

We acquired MRIs from 11 out of 14 stroke survivors and all 15 control participants. The remaining stroke survivors were not eligible for MRI. T1- and T2-weighted as well as multishell diffusion-weighted (DWI) images were acquired using a 3 T Prisma MRI scanner (Siemens Healthineers, Erlangen, Germany) with a 64-channel head coil. A 3-D magnetization-prepared rapid gradient echo sequence [repetition time (TR) = 2500 ms, echo time (TE) = 2.15 ms, flip angle 8°, 288 coronal slices with a voxel size of 0.8 × 0.8 × 0.8 mm³] was used. T2-weighted images were obtained with a fluid-attenuated inversion recovery sequence (TR = 9210 ms, TE = 92 ms, TI = 2500 ms, flip angle 140°, 70 axial slices with a voxel size of 0.9 × 0.9 × 2.0 mm³) for stroke lesion delineation. DWI was acquired by covering the whole brain with gradients (b = 500, 1000 and 2000 s mm^-2^) applied along 96 non-collinear directions with sequence parameters TR = 5000 ms, TE = 76 ms, slice thickness = 2 mm, in-plane resolution = 1 × 1 mm. For stroke survivor 14, the sequence parameters were as follows: TR = 5000 ms, TE = 78 ms, slice thickness = 2 mm, in-plane resolution = 1 × 1 mm.

### Data analysis

We used R version 4.1.3. and the software package Fieldtrip in MATLAB (R2021b, The MathWorks, Inc.).^29,30^

#### Clinical and movement data

To obtain ratio scores, whole hand grip force and key grip force scores of the performing hand were divided by scores of the non-performing hand. For ARAT, BBT and NHPT, the measure of the performing hand was used (ARAT: score with range 0 to 57; BBT: blocks/min, NHPT: pegs/sec). We used the absolute test scores for UEFM and MMSE (UEFM: range 0 to 66, MMSE: range 0 to 30). We defined *movement rate* as the reciprocal of the time between the first and the fourth button press in each trial, and *improvement* as the change in *movement rate* from the first block to the best block:

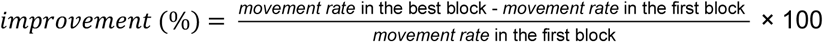

#### MEG Data

We bandpass filtered the MEG data between 1 and 120 Hz, and bandstop filtered between 49 and 51 Hz as well as between 99 and 101 Hz to eliminate the line noise artefact using a fourth-order butterworth filter. We then segmented the data into trials from −3.2 s to +3.5 s relative to the time point of the first button press. Artifacts were removed with a combination of visual artifact rejection and independent component analysis. We removed components representing eyeblinks, cardiac activity, or muscle artifacts. We excluded trials in which participants took more than 3 s to complete all button presses, trials in which the total movement duration (1^st^ to last button press) deviated more than three scaled absolute deviations from the individual block median, and trials with errors. This procedure left 188.9 ± 31.53 (mean ± standard deviation) trials per stroke survivor (2645 trials in total, stroke group) and 205.6 ± 14.6 trials per control participant (3084 trials in total, control group).

The dynamic imaging of coherent sources (DICS) beamforming method served to reconstruct source activity.^31^ We reconstructed individual head models from individual T1-weighted MRIs (11 stroke survivors, 15 control participants) or the Colin27 Average Brain (3 stroke survivors) with the single-shell method.^32,33^ Source models consisted of a regularly spaced 8 mm 3D grid warped on the individual or standard head model, respectively. For every participant, a projection matrix describing signal projection to the sensors, the leadfield matrix, was computed. Time-frequency averaged cross-spectral density matrices for the post-movement period (0.6 to 1.4 s after the last button press) and baseline period (−1.6 to −0.8 s before the first button press) at 21 ± 7 Hz were computed, and source space activity was estimated at each grid point. To noise-normalize source power, the difference of power between the movement period and the baseline period was divided by power in the baseline period for each trial. We identified brain regions by masking the source activity with the Brainnetome atlas.^34^ For the post movement period, source space activity was additionally estimated at 17 ± 3 Hz (“low beta”) and 25 ± 4 Hz (“high beta”).

For a time-frequency representation of characteristic activity, we additionally computed real-valued DICS filters at 21 ± 7 Hz for the periods −0.5 to 0.3 s around the first button press (task period), 0.6 to 1.4 s after the last button press (post-task period) and −1.6 to −0.8 s before the first button press (baseline period). We then multiplied the filters of the grid points with maximal power in each region with the time-series data. In the resulting source level time-series data, we estimated power at 21 ± 7 Hz. MEG data of participants conducting the task with their right hand were mirrored across the hemispheres.

#### MRI-Data: Fractional anisotropy of the corticospinal tract

We used QSIPrep 0.16.1, based on Nipype 1.8.5 for preprocessing and reconstruction of MRIs.^35,36^ Segmentation of the corticospinal tract (CST) was done using TractSeg.^37^ Tract-related mean fractional anisotropy (FA) values of corresponding DWI voxels were extracted using MRtrix3.^38^ Mean CST FA values from the hemisphere contralateral to the performing hand (the affected hemisphere in stroke survivors) were divided by those from the ipsilateral hemisphere to compute ratio scores.

### Data analysis and statistics

#### Group differences in clinical and behavioural data

We assessed differences in clinical characteristics, *improvement* and average block 1 *movement rate* with two-tailed *t*-tests and Wilcoxon rank-sum tests as appropriate.

#### Beta ERS and ERD

We calculated paired two-tailed *t*-tests for each source grid point between the post movement period (0.6 to 1.4 s after the last button press), the movement period (−0.5 to 0.3 s around the first button press), and the baseline (−0.8 to −1.6 s before the first button press). Beta power in the post movement period (ERS) and movement period (ERD) were tested for significance against the baseline period via cluster-based nonparametric statistical tests with 10000 randomisations and a cluster threshold of *α* = 0.025.^39^ Similarly, we tested for differences in beta ERS between stroke survivors and control participants. Additionally, the same analyses were conducted for low (14 – 20 Hz) and high (21 – 29 Hz) beta ERS power.

#### Relationship of beta power with improvement and movement rate

For each group, we calculated linear models to examine the relationship between *improvement* or mean *movement rate* and beta power (either beta ERS or ERD) across participants. The performing hand was included as a fixed effect in all models. Additionally, in the model assessing the association of *improvement* and beta power, *movement rate* in block 1 was included as a fixed effect to account for potential scaling of *improvement* based on baseline performance levels.

These linear models were computed for activity at each grid point in source space to assess the cortical topography of the relation between beta power and the variable of interest (*improvement* or mean *movement rate)*. For circumvention of the problem of multiple comparisons, we applied a cluster-permutation analysis with 1000 randomisations of the assignment between beta power and the variable of interest while keeping the assignment of the controlling factors group and performing hand to beta power fixed.^39^ The cluster threshold was set to *α* = 0.01. We compared the summed t-values of the variable of interest within each cluster to the distribution of summed t-values within clusters after randomisation to compute the final p-value of the cluster.

For the detection of possible differences across the cohorts in how beta power and motor performance relate, we furthermore conducted an analysis of the model including data of both cohorts and an interaction of *improvement* or *movement rate* and group. Again, this analysis was done for all grid points in source space and a cluster-permutation analysis determined whether significant clusters of the interaction were found.

Furthermore, we detected the maximum ERS of every participant within a significant cluster (“cluster ERS”) and utilized it to recalculate the linear model in a simpler, representative form (*cluster ERS ∼ performed hand + movement rate block 1 + improvement x group*). For this representative model, we checked the general assumptions for linear models (linearity, absence of collinearity, homoscedasticity, normality of residuals, absence of influential observations) and conducted a leave-one-out analysis.

#### Dependence of the relationship between beta power and improvement on lesion location and beta frequency band

Additionally, we calculated the model including stroke survivors with only subcortical lesions as well as with only cortical lesions. To investigate whether the model results are determined by low or high beta power, we again extracted the maximum ERS of every participant within the cluster for low frequency (14-20 Hz) and high frequency (21-29 Hz) beta power and re-computed the linear model for both frequency ranges.

#### Exploratory analysis of factors influencing the relationship of beta power and improvement

To find out which physiological and structural factors may influence the relationship of beta ERS and motor skill acquisition, we added further exploratory analyses of the cluster ERS model. The model was analogous to the main model, with performing hand and movement rate in block 1 as fixed effects. We tested for the interactions of *improvement* x group with UEFM, ARAT, BBT, NHPT, grip force, key grip force, CST FA ratio, and age in relating to the cluster ERS. All factors were tested separately. Results of this exploratory analysis were not corrected for multiple comparisons.

## Results

### Participants

The study involved 14 stroke survivors (mean age 65.4 years, SD = 9.3; 7 female participants; 8 with right hemisphere lesions; performing hand: 8 left, 6 right) and a control group of 15 age- and sex-matched individuals (mean age 64.5 years, SD = 8.4; 7 female participants; hand used for task: 10 left, 5 right). None of the participants exhibited cognitive impairment based on MMSE scores (stroke group: mean score 28.9, range 27–30; control group: mean score 29.7, range 28–30).

### Clinical scores and structural imaging

Stroke survivors exhibited mild upper limb impairment, as reflected by a mean UEFM of 61.1 (Table 1). As would be expected, significant group differences were observed between stroke survivors and control participants for UEFM, ARAT, BBT and NHPT measures (Fig. 1B, C; UEFM: *W* = 30.5, *p* < 0.001, ARAT: *W* = 51.0, *p* = 0.004, BBT: *t*_19.2_ = −3.8, *p* = 0.001, NHPT: *t*_18.9_ = −2.7, *p* = 0.016). In contrast, no significant differences were found for whole hand grip force ratio (Fig. 1D; *W* = 64.0, *p* = 0.077), or key grip force ratio (Fig. 1D; *t*_15.8_ = 0.4, *p* = 0.688). Notably, CST FA ratios overlapped substantially between groups, showing no significant difference (Fig. 1E; *t*_22.8_ = 1.1, *p* = 0.269), indicating comparable corticospinal tract integrity across participants.

**Figure 1.**
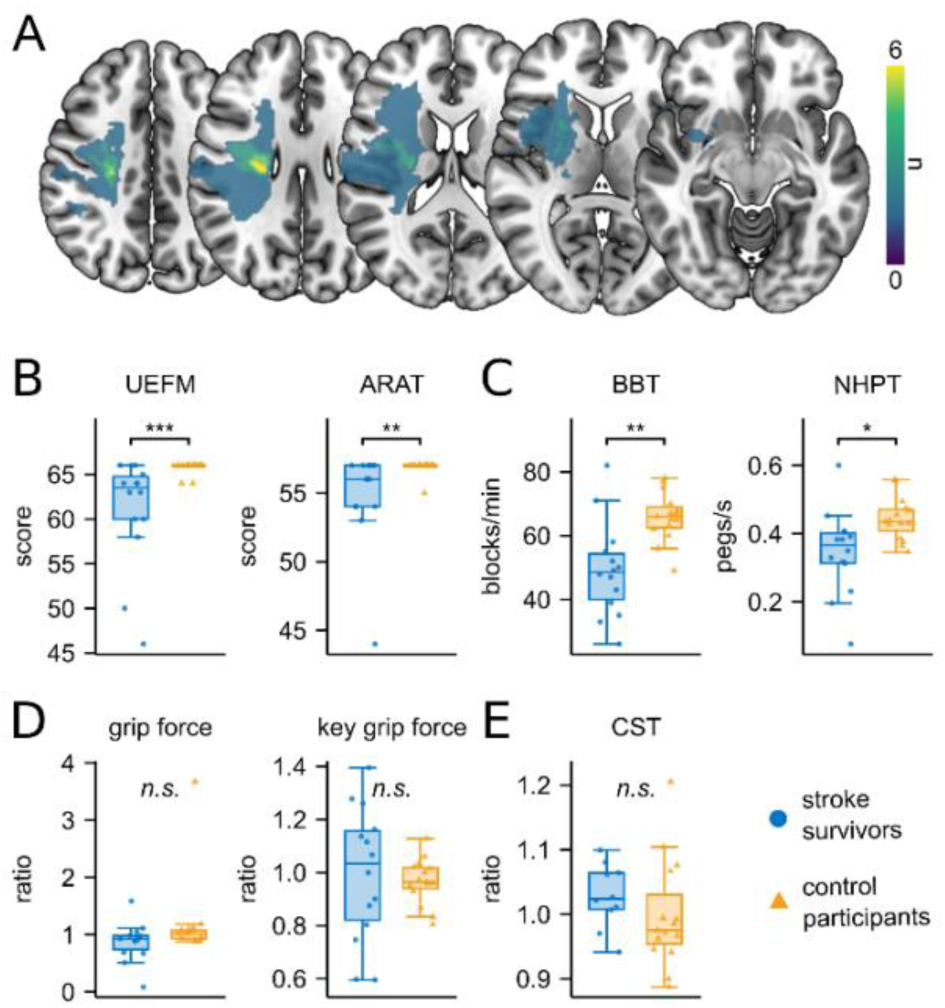
Clinical stroke characteristics. (**A**) Lesions of the 10 stroke survivors with available MRI lesion data, overlayed on an MNI template. Colour indicates the number of individuals with a lesion at this voxel. Left-hemispheric lesions were flipped to the right hemisphere. (**B-D**) Clinical scores. Tested for group differences, no correction for multiple comparisons. Significance markers: \**p* < 0.05, \*\**p* < 0.01, \*\*\**p* < 0.001. (**B**) Absolut UEFM scores and absolute ARAT score of the affected hand (stroke survivors) or performing hand in motor task (control participants). (**C**) Values of Box and Block Test (BBT, blocks/min) and Nine Hole Peg Test (NHPT, pegs/s) of the affected hand (stroke survivors) or performing hand (control participants). (**D**) Whole hand grip force and key grip force: Ratios of performing hand to non-performing hand. (**E**) Distribution of CST-FA ratios.

**Table 1.** Demographic and clinical data of stroke survivors.

| Stroke survivor | Sex | Age Group | Lesion side | TAS (months) | Stroke location | MRS | NIHSS | UEFM | ARAT | Grip force | KG force | NHPT | BBT |
| --- | --- | --- | --- | --- | --- | --- | --- | --- | --- | --- | --- | --- | --- |
| 1 | F | 70-79 | L | 52 | TC, CR | 1 | 0 | 65 | 57 | 1.11 | 1.16 | 0.39 | 50 |
| 2 | F | 70-79 | R | 8 | TC, CR, CI | 3 | 2 | 46 | 44 | 0.09 | 0.60 | 0.08 | 26 |
| 3 | M | 70-79 | R | 153 | BG, CI, CR | 2 | 1 | 50 | 54 | 0.51 | 0.60 | 0.20 | 33 |
| 4 | F | 70-79 | R | 52 | BG | 1 | 1 | 63 | 57 | 0.97 | 1.07 | 0.35 | 43 |
| 5 | F | 70-79 | R | 43 | PLCI | 1 | 0 | 60 | 56 | 0.89 | 0.80 | 0.38 | 47 |
| 6 | F | 40-49 | L | 38 | PLCI | 0 | 0 | 66 | 57 | 1.59 | 1.26 | 0.60 | 82 |
| 7 | F | 70-79 | L | 39 | MED | 1 | 1 | 58 | 56 | 0.92 | 0.88 | 0.31 | 49 |
| 8 | M | 60-69 | L | 9 | BG | 1 | 0 | 66 | 57 | 0.93 | 1.28 | 0.45 | 71 |
| 9 | M | 60-69 | R | 9 | PLCI | 1 | 0 | 66 | 56 | 0.89 | 1.12 | 0.32 | 39 |
| 10 | M | 50-59 | L | 6 | CR | 1 | 0 | 64 | 57 | 1.05 | 1.00 | 0.40 | 52 |
| 11 | M | 50-59 | L | 51 | MED | 1 | 2 | 63 | 53 | 0.98 | 1.14 | 0.38 | 55 |
| 12 | F | 60-69 | R | 129 | PO | 0 | 0 | 64 | 57 | 0.97 | 1.40 | 0.40 | 58 |
| 13 | M | 60-69 | R | 9 | CR | 1 | 1 | 60 | 54 | 0.68 | 0.90 | 0.23 | 35 |
| 14 | M | 60-69 | R | 16 | PRE | 1 | 0 | 64 | 54 | 0.69 | 0.75 | 0.33 | 48 |
| Mean stroke | F: 7 | 65.4 |  | 45.6 |  | 1.1 | 0.6 | 61.1*** | 54.93** | 0.88 | 1.00 | 0.35* | 49.14** |
| SD stroke |  | 9.3 |  | 45.3 |  | 0.7 | 0.8 | 6.1 | 3.45 | 0.33 | 0.25 | 0.12 | 14.73 |
| Mean control | F: 7 | 64.5 |  |  |  | 0 | 0 | 65.7 | 56.87 | 1.18 | 0.97 | 0.44 | 65.87 |
| SD control |  | 8.4 |  |  |  | 0 | 0 | 0.7 | 0.52 | 0.7 | 0.09 | 0.06 | 7.65 |

\* $p < 0.05$ , \*\* $p < 0.01$ , \*\*\* $p < 0.001$ , statistical test of stroke vs control group, uncorrected. Values of grip force and key grip (KG) force are given as ratios of the affected to the non-affected hand in stroke survivors, and of the performing to the non-performing hand in control participants. Values of MRS, NIHSS, ARAT, NHPT (pegs/s) and BBT (blocks/min) are reported for the affected hand in stroke survivors and performing hand for control participants. UEFM is stated as the absolute score for both groups. TAS: time after stroke, SD: standard deviation, TC: thalamocapsular, CR: corona radiata, CI: capsula interna, BG: basal ganglia, PLCI: posterior limb of the capsula interna, MED: media infarct, PO: pons, PRE: precentral gyrus

#### *Improvement* in *movement rate* is comparable in stroke survivors and control participants

Both stroke survivors and control participants demonstrated *improvement* in *movement rate* throughout the task: From block 1 to block 6, both groups showed an increase in *movement rate* (Fig. 2B left column, stroke survivors: *t*_12.0_ = −4.7, *p_cor_* = 0.001, control participants: *t*_14.0_ = - 6.9, *p_cor_* < 0.0001, corrected for two comparisons). Nevertheless, on average, stroke survivors had a significantly lower initial *movement rate* in block 1 than control participants (Fig. 2B right column, *t*_21.3_ = −3.9, *p* < 0.001). When comparing the initial block to the individual best block, stroke survivors showed an *improvement* of 22.6 ± 12.2% (mean ± standard deviation), while control participants improved by 15.3 ± 7.1%. Even though there was a trend towards lower *improvement* in the control group, the extent of *improvement* did not significantly differ between groups (Fig. 2C, *t*_20.5_ = 1.9, *p* = 0.066).

**Figure 2.**
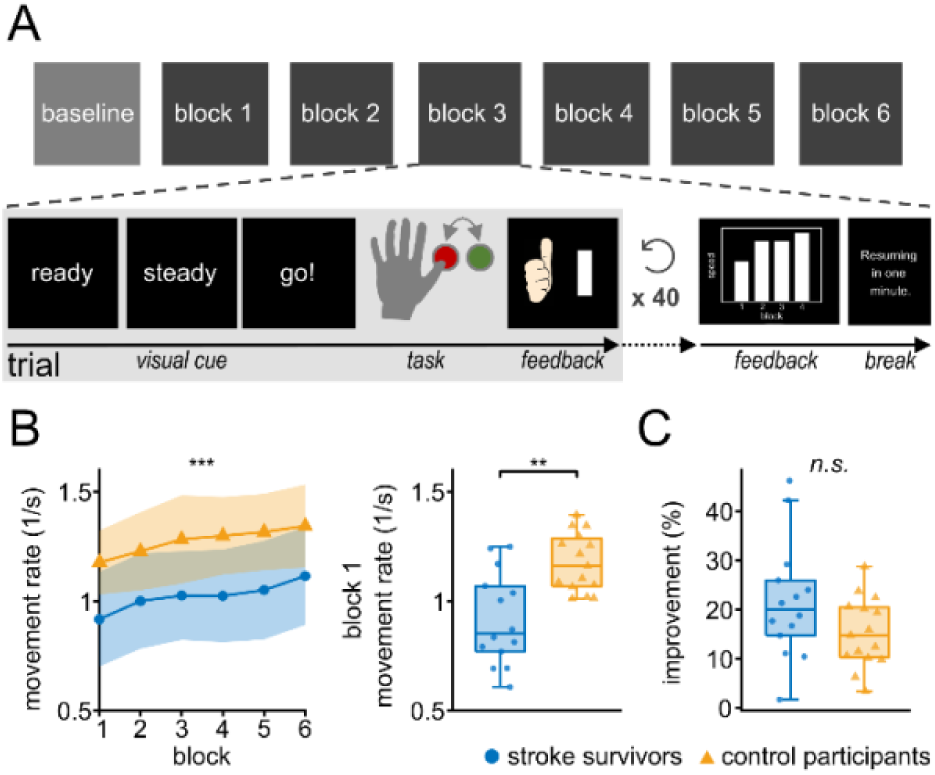
Motor skill acquisition task and summary of movement data. (**A**) The task consisted of one baseline block of 20 trials and six measurement blocks, each containing 40 trials. During each trial, participants performed four alternating button presses in maximum speed in response to a visual cue. Participants received real-time feedback on their *movement rate* after each trial, as well as additional feedback showing the block-averaged *movement rate* after each block. Between blocks, participants were given a one-minute rest period. (**B**) Left: The *movement rate* for each trial was averaged within each group, revealing a consistent increase over the course of the trials in both groups. Shaded areas indicate ± 1 standard deviation. Right: Distribution of participant mean block 1 *movement rate*. Asterisks indicate significant group differences. Significance markers: \*\**p* < 0.01, \*\*\**p* <0.001 (**C**) Distribution of *improvement* for all participants. *Improvement* is defined as the percentage increase in *movement rate* from the first to the individual’s best block. There were no significant group differences in *improvement*.

### Movement-related beta power in stroke survivors and control participants

First, we confirmed typical characteristics of beta power modulation around movement. As expected, we observed a cluster with a significant decrease in beta power (14 – 28 Hz) during the movement period relative to baseline (−0.5 to 0.3 s around the first button press), known as beta ERD, centred over sensorimotor cortices (stroke survivors: *p_cor_* < 0.0001, control participants: *p_cor_* < 0.0001; corrected for two comparisons, Fig 3A, C). Following the movement period (0.6 to 1.4 s after the last button press), beta power increased relative to baseline, reflecting beta ERS (stroke survivors: *p_cor_*< 0.0001, control participants: *p_cor_* < 0.0001; corrected for two comparisons, Fig. 3B, D). A permutation-based cluster statistics across the whole brain, conducted without any prior assumptions on regions of interest, did not identify significant clusters when comparing beta ERD and ERS between groups (ERS: *p* = 0.16, ERD: *p* = 0.11, not corrected for multiple comparisons, Fig. 3E, F), although there was a trend toward lower beta ERD and ERS in stroke survivors. However, in the ipsilesional ventral premotor cortex (PMv), contralateral to the movement, beta ERS was significantly lower in stroke survivors than control participants, while the ERD showed no group difference (ERD: *t*_21.6_ = 1.6, *p_cor_* = 0.26; ERS: *W* = 52.0, *p_cor_* = 0.04; corrected for two comparisons, Fig. 3E, F). Similar to the wide-range beta ERS, group comparisons of low (14 – 20 Hz) and high (21 – 29 Hz) beta ERS revealed non-significant clusters, with a trend toward reduced power in stroke survivors (low beta ERS: *p* = 0.13, high beta ERS: *p* = 0.15, not corrected for multiple comparisons, Suppl. Fig 1B). Within each group, ERS power was significantly higher in the low beta band compared to the high beta band in a cluster spanning wide areas of cortex (stroke survivors: *p_cor_* < 0.001, control participants: *p_cor_* < 0.001, corrected for two comparisons, Suppl. Fig. 1C).

**Figure 3.**
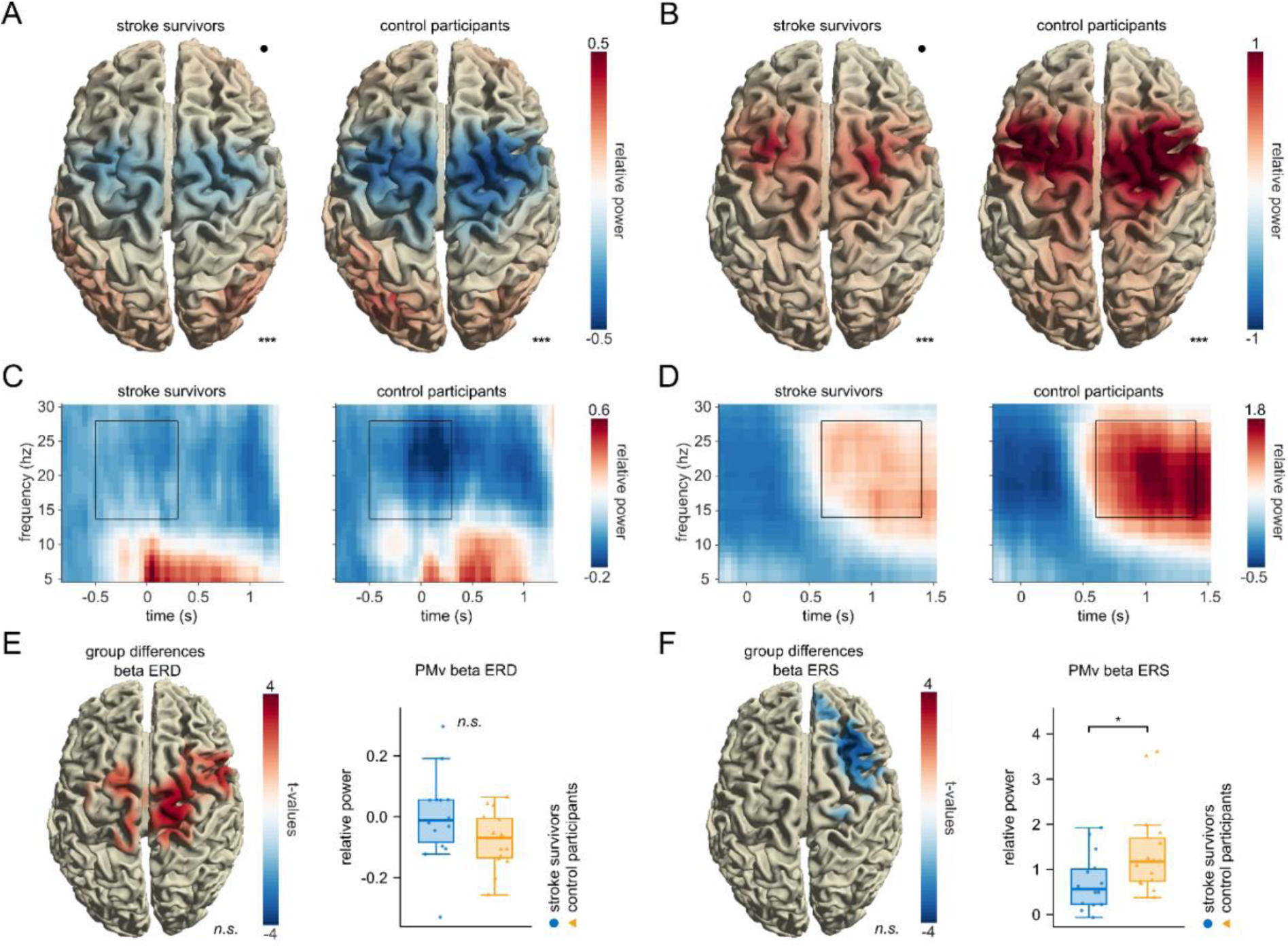
Beta ERD and ERS. (**A**) Group-mean beta ERD and **(B)** beta ERS projected onto a standard brain. The colour represents the change in beta power relative to baseline. The black dot represents the lesion side (right hemisphere). For stroke survivors with lesions on the left side and for participants who performed the task with the right hand, activity was mirrored across hemispheres. The time-frequency window is selected as depicted in (**C**) and (**D**). (**C**) and **(D)** Time-frequency representation in PMv for stroke survivors and control participants. The selected time window is marked as a black rectangle (14 – 28 Hz, ERS: 0.6 – 1.4 s after the last button press, ERD: −0.5 – 0.3 s around the first button press). (**E**) and (**F**) Results of whole-brain permutation-based cluster statistics comparing beta ERD (**E**, left) and ERS (**F**, left) power between stroke survivors and control participants. No significant clusters were found, while there was a trend toward a lower ERD and lower ERS in stroke survivors compared to control participants (ERD: *p* = 0.09, ERS: *p* = 0.17, not corrected for multiple comparisons). The beta ERS in the ipsilesional ventral premotor cortex (PMv) was significantly reduced in stroke survivors compared to control participants (**F**, right), whereas the difference in ERD (**E**, right) was not significant (ERD: *t*21.6 = 1.6, *pcor* = 0.26; ERS: *W* = 52.0, *pcor* = 0.04; corrected for two comparisons). Significance markers: *n.s.* = not significant, \**p* < 0.05, \*\*\**p* < 0.001.

### Beta ERS relates positively to *improvement* in control participants, but not in stroke survivors

Next, we investigated how beta ERD and ERS were related to *movement rate* and *improvement*. We applied linear models to assess the relationship between beta power and both mean *movement rate* and *improvement* at each grid point. Analyses were conducted separately for each group to identify group-specific associations, and additionally, for the group x *improvement* interaction to determine whether the relationship between beta power and behaviour differed between stroke survivors and control participants. Of particular interest was the association between beta ERS and *improvement* (Fig. 4). In stroke survivors, no significant clusters linking beta ERS and *improvement* were found (Fig. 4A). However, in control participants, greater *improvement* was significantly associated with higher beta ERS in a cluster spanning the bilateral sensorimotor cortices (*p* < 0.001, cluster statistic: 5.3 × 10³, SD < 0.001, CI: 0.002, Fig. 4B). These results are further supported by the interaction model, which revealed a significant group x *improvement* interaction (*p* = 0.002, cluster statistic: 3.67 × 10³, SD = 0.001, CI: 0.003, Fig. 4C). Selecting the grid point with maximal beta ERS power for each participant within the cluster (“cluster ERS”) confirmed the different association between *improvement* and beta ERS in stroke survivors and control participants (overall regression: adjusted R² = 0.63, F statistics = 10.51, *p* < 0.0001; *improvement* x group interaction term: β = −0.18, *p* < 0.00001; Fig. 4D) with a trend towards a negative association in stroke survivors. Notably, the relation of beta power and *improvement* was specific for beta ERS and was not observed for beta ERD (Suppl. Fig. 2). Additionally, neither beta ERS nor beta ERD related significantly to mean *movement rate* (Suppl. Fig. 3).

**Figure 4.**
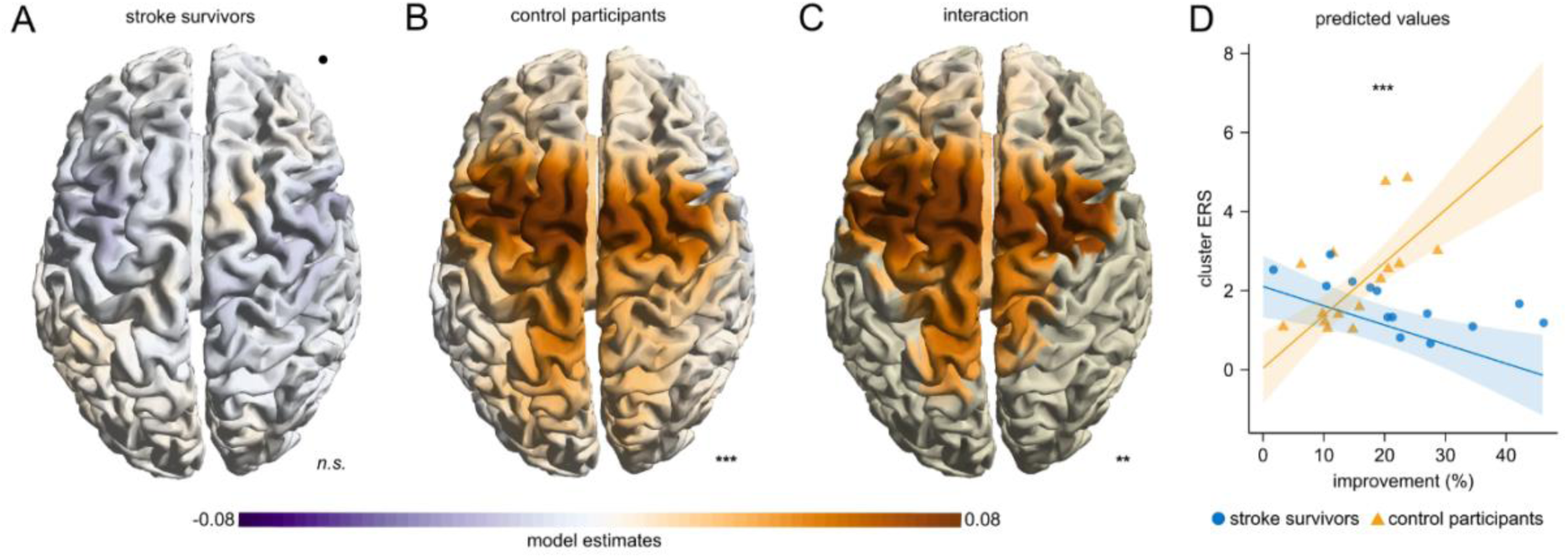
Beta ERS relates to *improvement* in control participants. (**A, B**) Grid point-wise linear model of *improvement* relating to the ERS. Model estimates for the factor *improvement* are plotted. A significant cluster in control participants (**B**) but not in stroke survivors (**A**) was found. The black dot represents the lesion side (right hemisphere). (**C**) Grid point-wise linear model, showing model estimates for the interaction of group x *improvement* relating to the ERS, including data of both stroke survivors and control participants (*p* = 0.002). The significant cluster was masked. (**D**) Linear model showing the interaction of group x *improvement* relating to the cluster ERS. Participant data points are shown as circles (stroke survivors) and triangles (control participants).

To validate our model, we tested standard assumptions of linear models using the representative model relating to cluster ERS (Figure 4D). Visual inspection of residual plots did not reveal any obvious deviations from homoscedasticity or normality. Data of two participants (14 and 24, one stroke survivor and one control participant) were identified as influential observations. However, the model still showed a robust interaction when leaving out these two participants (*improvement* x group interaction term: *p* < 0.0001). Leaving out any single participant out of all 29 participants also revealed robust interactions without exception (*improvement* x group interaction term: all *p* < 0.0001).

### Results do not depend on lesion location or beta sub-band

Furthermore, we investigated if our results may depend on the lesion location or the exact beta frequency band. Computing the model with a subgroup of stroke survivors with solely subcortical (n=11) as well as with cortical lesions (n=3) consistently led to a significant group x *improvement* interaction (Figure 5A, only stroke survivors with subcortical lesions included: overall regression: adjusted R² = 0.63, F statistics = 9.45, *p* < 0.0001, *improvement* x group interaction term: β = −0.19, *p* < 0.0001; Figure 5B, only stroke survivors with cortical lesions included: overall regression: adjusted R² = 0.67, F statistics = 7.91, *p =* 0.002, *improvement* x group interaction term: β = −0.19, *p* < 0.001). Moreover, model results remained consistent when using the maximum cluster grid point of either low (14-20 Hz) or high (21-29 Hz) frequency beta power instead of the chosen wide range beta (Figure 5C, low frequency beta ERS: overall regression: adjusted R² = 0.67, F statistics = 12.12, *p* < 0.00001, *improvement* x group interaction term: β = −0.31, *p* < 0.00001; Figure 5D, high frequency beta ERS: overall regression: adjusted R² = 0.53, F statistics = 7.38, *p* < 0.001, *improvement* x group interaction term: β = −0.15, *p* < 0.0001).

**Figure 5.**
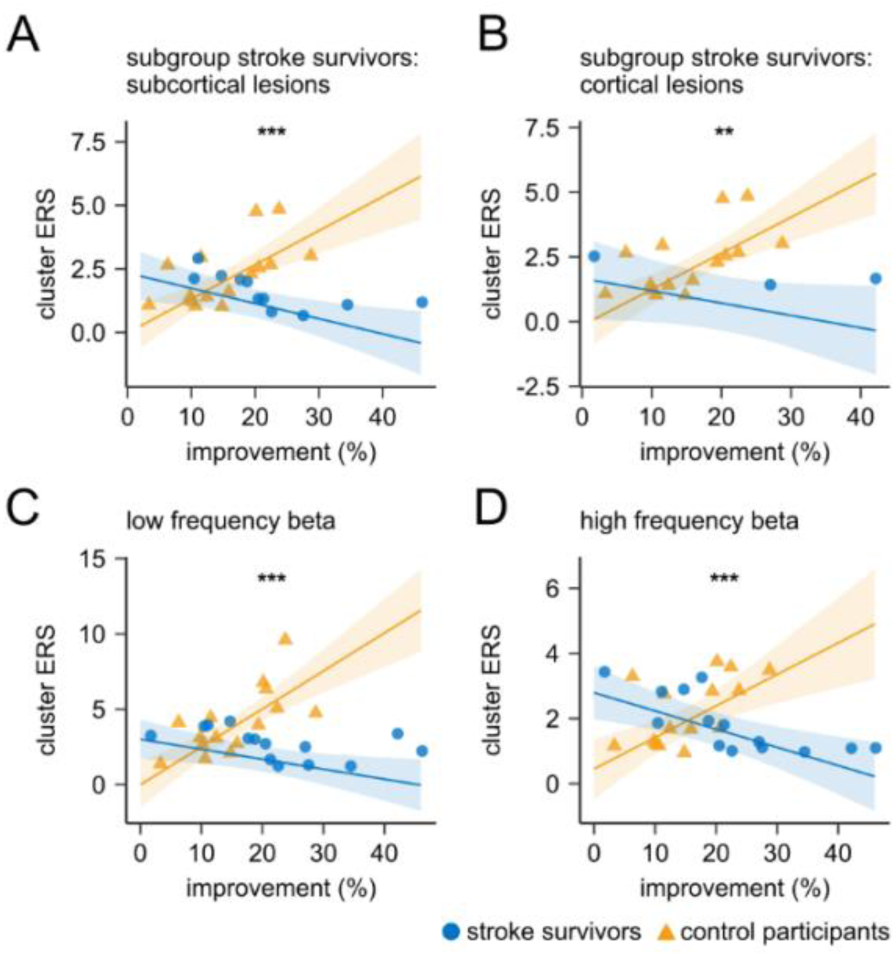
Dependence of the group-specific relationship of ERS and *improvement* on lesion location and beta frequency band. Main model (“cluster ERS”) calculated with only stroke survivors with (**A**) subcortical and (**B**) cortical lesions. Main model also calculated for maximum beta power within the cluster for (**C**) low frequency beta and (**D**) high frequency beta.

#### Influence of clinical scores and CST integrity on beta ERS–*improvement* relationship

Given the observed differences in the relationship between beta ERS and *improvement* across groups, we conducted an exploratory analysis to assess whether clinical scores, age or CST integrity influenced this association. For each factor, we modelled the interaction between this factor, *improvement* and group to be associated with the cluster ERS. Only the model including the BBT, assessing manual dexterity and gross motor function, reached significance for this triple interaction (p = 0.016). In contrast, other clinical tests of arm movements, daily activities, and fine motor coordination showed no significant triple interaction (Supplementary Table 2; all p > 0.05). Specifically, calculating the model for each group separately revealed an enhanced relationship between *improvement* and beta ERS power in control participants with higher BBT, while no significant influence of BBT could be seen in stroke survivors (Fig. 6, stroke survivors: overall regression: adjusted R² = 0.67, F statistics = 6.18, *p* = 0.01, *improvement* x BBT interaction term: β = 0.0002, *p* = 0.81; control participants: overall regression: adjusted R² = 0.79, F statistics = 11.82, *p* < 0.001, *improvement* x BBT interaction term: β = 0.01, *p* = 0.04).

**Figure 6.**
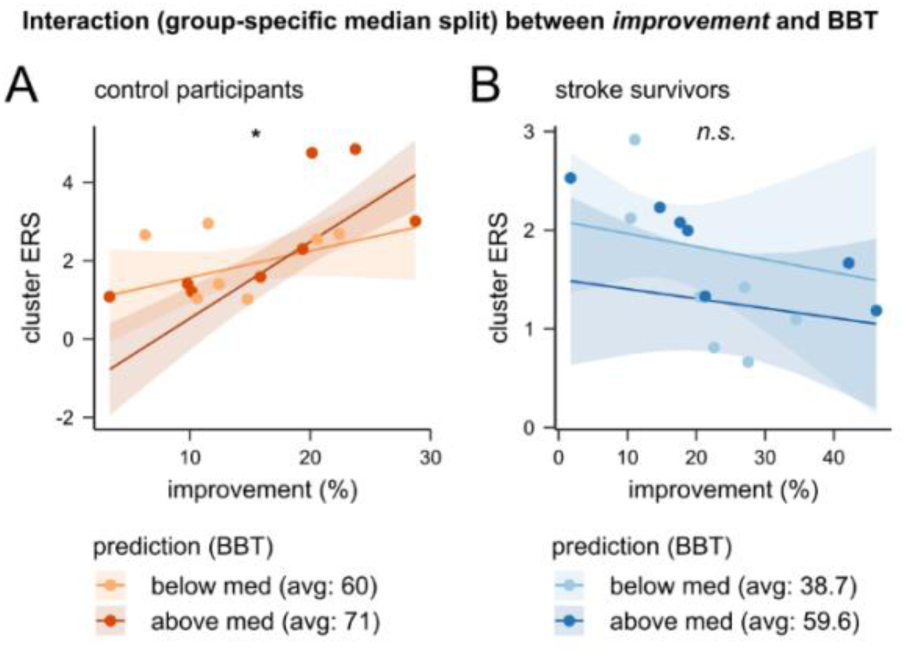
Relationship between cluster ERS, group, *improvement*, and BBT score. Separate linear models for control participants (**A**) and stroke survivors (**B**) with a BBT score median split to illustrate how the interaction between *improvement* and BBT score relates to beta ERS power.

## Discussion

Diverse roles have been associated with beta activity during and after movement but are still in the process of being defined. Here, we investigated the function of beta ERD and beta ERS for unimanual motor skill acquisition in stroke survivors and control participants using MEG source-reconstructed activity. We found that, while beta ERS strongly related to motor skill acquisition in control participants, this relationship was absent after stroke. Furthermore, no association of beta ERD and motor skill acquisition was observed. The main finding, an interaction of *improvement* (quantifying motor skill acquisition) and group associated with beta ERS, proved to be statistically robust, mainly related to the strong association of beta ERS and *improvement* in the control cohort. In particular, the interaction remained strong regardless of stroke location (subcortical vs cortical) or specific beta frequency bands.

### Topological organization of the relationship between beta ERS and *improvement*

Both ERD and ERS occurred bilaterally in sensorimotor areas consistent with previous literature (Fig. 3).^5,40^ In line with this location, also the association of *improvement* with beta ERS in control participants, and the interaction of *improvement* and group across the cohorts was found bilaterally, even with stronger pronunciation of the hemisphere ipsilateral to the performing hand (Fig. 4). In contrast, but in accordance with earlier studies, differences in beta ERS power across the cohorts were rather located on the hemisphere contralateral to the performing hand (Fig. 3F).^23^ This difference in location suggests that the potentially reduced beta ERS after stroke and our main finding – differences across cohorts in the relationship of beta ERS with motor skill acquisition – are separate findings that may not necessarily depend on each other.

### Roles of beta ERS for motor control and motor skill acquisition

The connection of beta ERS and motor skill acquisition in healthy participants is not a new finding.^7–11^ For example, Tatti et al.^11^ described that beta modulation, the difference of ERD and ERS, is stronger during a learning condition than during a mere motor task.^11^ Espenhahn et al.^7^ found that beta ERS is associated with motor performance one hour after a motor skill acquisition task. Besides motor skill acquisition, a relation of beta ERS with movement error has been described.^12–14,41^ Tan et al.^41^ suggested beta ERS as a marker of the uncertainty of the inner model predicting movement execution. Following this theory, higher beta ERS is associated with less uncertainty in predicting movements.

This theory^41^ could align with our findings in healthy adults: those who improve more may do so because their internal model becomes more certain, reducing error and thus enhancing beta ERS associations with motor skill acquisition. In stroke survivors, by contrast, this linkage may be disrupted, or skill acquisition may proceed without parallel increases in predictive certainty. In support of this interpretation, Korka et al.^8^ demonstrated that beta ERS is also sensitive to the cognitive demands of strategy learning, with greater decreases of the ERS when participants had to acquire and apply a re-aiming strategy. Their results suggest that beta activity reflects not only prediction error and uncertainty, but also the engagement of higher-order cognitive processes during motor learning.

In contrast to the studies with healthy participants, few studies have investigated motor skill acquisition and beta ERS after stroke. Specifically, Espenhahn et al.^23^ found that beta ERS of stroke survivors after training correlated with better task performance later on and thus motor skill acquisition. In contrast to our study, beta ERS after training was related to the skill 24 hours post-training, thus referring to motor maintenance instead of short-term motor skill acquisition. Moreover, the stroke survivors in the study of Espenhahn et al.^23^ showed significantly lower motor skill acquisition compared to control participants, which was not the case in our study. Tang et al.^20^ did not investigate motor skill acquisition but motor recovery and showed an association of beta ERS with increased motor function (UEFM) twelve weeks post-stroke using source-space MEG. Our study differs from these two studies with respect to the largely different time scales of investigation, and our cluster-based approach in MEG source space.

Furthermore, in our study, beta ERS did not scale with *movement rate* (quantifying task performance), while in previous reports, beta ERS has been related to motor function.^15,17,20,42,43^ One possible reason for this discrepancy is that performance in our task is not suitable to evaluate general motor function as the task complexity was relatively high, requiring both speed and coordination, leading to consistently strong task-related activity. For our stroke cohort, it is possible that a direct association between beta ERS and motor function may not be detected as the stroke survivors were rather mildly affected (NIHSS 0-2), whereas in most previous studies the range of impairment was wider. Nevertheless, Laaksonen et al.^15^ reported an association of NHPT results with beta ERS also at levels of low impairment, while the strength of this association was weak.

Previously, a reduction of beta ERS after stroke has been reported.^16,19,20^ We also found this reduction locally in pMV, but not when calculating cluster statistics on the whole brain (Fig. 3F). Possibly, our whole-brain cluster statistics, indirectly taking multiple comparisons into account, were not sensitive enough to detect this local reduction. Another explanation is again that our cohort consisted of well-recovered stroke survivors, as suggested by Espenhahn et al.^23^, who similarly found no beta ERS differences between mildly affected stroke survivors and healthy control participants prior to training. Nevertheless, Kulasingham et al.^19^ reported reduced beta ERS even in participants without severe impairments. This discrepancy may possibly relate to differences in task demands. Specifically, our high task complexity may have recruited a wider network of motor-related beta sources, potentially compensating for localized deficits and thereby masking group-level differences in beta ERS.

Moreover, accumulating evidence suggests that low- and high-beta activity may subserve distinct functions in motor control.^44^ In our study, the relationship between beta ERS and *improvement* was comparable for low and high beta ERS (Fig 5C, D). However, we observed that low beta ERS was consistently and significantly stronger than high beta ERS in both stroke survivors and healthy control participants (Supplementary Fig. 1C). The absence of differential behavioral associations in our dataset may indicate that both sub-bands contribute jointly to post-movement dynamics, but with low beta activity exerting a dominant influence on the overall ERS profile.

### Mediators of the relationship between motor skill acquisition and beta ERS

CST damage has been shown to be one major predictor of clinical recovery after stroke.^45^ Furthermore, beta activity has been associated with CST damage,^46^ and the association of movement-related beta activity and impairment was driven by the extent of CST damage.^47^ In contrast, in our study, the interaction of *improvement* and group was statistically not related to CST damage. However, the CST of the stroke survivors in this study was only partially damaged or indeed was intact. Furthermore, only the stroke survivors with MRI (11 out of 14) could be assigned CST FA ratio values, restricting statistical power. Instead, the interaction was related to the BBT score, representing gross manual dexterity and focusing on speed and coordination. Specifically, the association of *improvement* and beta ERS in control participants depended on the BBT score, with lower BBT values (lower gross manual dexterity) coming along with a weaker association. For stroke survivors, who generally showed lower BBT scores, the association of *improvement* and beta ERS was broken. While those analyses had an exploratory character and need to be confirmed independently, they suggest that the critical difference between the two groups may not be the stroke per se, but rather the motor function of participants. In other words, beta ERS may support motor skill acquisition when gross manual dexterity is already at a high level, but other neural mechanisms could maintain motor skill acquisition when gross manual dexterity is suboptimal.

### Roles of beta ERD and other neural dynamics for motor skill acquisition

In line with previous studies,^13,48,49^ we did not observe significant relations of beta ERD with motor skill acquisition, and we also did not find beta ERD to relate to performance. Beyond beta-band activity, oscillations in other frequency ranges could contribute to motor skill acquisition. Most prominently, gamma-band activity (25-140 Hz) is thought to reflect local circuit plasticity and has been linked to the consolidation of newly acquired motor patterns.^50^ In contrast, in our previous work, also high-gamma was linked to motor skill acquisition neither in stroke survivors nor control participants.^27^ Similarly, Wu et al.^13^ did not find an association of gamma activity with movement learning or error in healthy participants. Together, these results suggest potentially distinct contributions of beta ERS and other neural activity within the motor system.

### Considerations

Several considerations are important for our study. First, the sample size of stroke survivors (n = 14) and control participants (n = 15) was restricted, making it possible that we missed findings of smaller effect size because of restricted statistical power. Despite the small sample size, we showed strong associations of beta ERS with *improvement* in control participants. The statistical power may be enhanced by our methods, which aggregate the spatially resolved MEG data into clusters without making a priori assumptions on the location. In addition, most stroke survivors had only mild motor impairment. Thus, it is not clear to which extent our results would be generalizable also to stroke survivors with stronger deficits. Second, the group of stroke survivors was relatively heterogeneous, including both cortical and subcortical strokes with a dominance for subcortical strokes. However, in secondary analyses including only cortical or only subcortical strokes, our main finding was confirmed in both subgroups. Potentially, the main result, the interaction of *improvement* and group, is driven by the strong relationship of beta ERS and *improvement* in the control cohort. Finally, as we investigated isolated thumb movements, the generalisability to other movements of daily life is unknown.

### Clinical implications

Our findings suggest that beta ERS in sensorimotor cortical areas may not serve as a reliable biomarker for short-term motor skill acquisition after stroke. Furthermore, interventions aimed at enhancing beta ERS, such as non-invasive neuromodulation techniques, may not necessarily lead to immediate improvements in motor skill acquisition. However, previous studies have reported associations between beta ERS and long-term motor recovery,^20,23^ indicating that beta ERS may play a more prominent role in sustained functional improvements rather than in rapid skill acquisition. This raises the question whether modulating beta ERS through approaches like tACS could support long-term recovery processes in stroke survivors, even if short-term learning remains unaffected. Future interventional studies that directly manipulate beta ERS and assess both short- and long-term motor outcomes are needed to clarify its causal role in post-stroke recovery and to determine whether beta ERS is a viable therapeutic target in neurorehabilitation.

## Conclusion

Our results indicate that beta oscillations play multiple roles in motor skill acquisition, and that these roles may shift in the presence of motor impairment or reduced gross manual dexterity. These findings have important implications for post-stroke neuromodulation strategies: enhancing beta ERS may not uniformly improve short-term motor skill acquisition, particularly in heterogeneous stroke populations. Tailoring neuromodulatory interventions to individual motor profiles may therefore be crucial to optimizing outcomes in neurorehabilitation.

## Data Availability

Data and code for this study are available via GitLab (https://gitlab.rrz.uni-hamburg.de/xeni/beta-synchronisation-after-stroke).

## Acknowledgements

We are thankful to Karin Reimann and Christiane Reißmann for assistance in data recording and recruitment of participants, and Kirstin-Friederike Heise as well as Sophie Grigutsch for helpful discussions.

## Author contributions

L.S.T.: Conceptualization, Data curation, Formal analysis, Investigation, Methodology, Software, Validation, Visualization, Writing – Original draft. B.H.: Conceptualization, Investigation, Methodology, Software, Writing – Review & Editing. S.W.: Investigation, Writing – Review & Editing. C.J.S.: Conceptualization, Writing – Review & Editing. J.F.: Software, Writing – Review & Editing. T.V.L.: Methodology, Writing – Review & Editing. R.S.: Methodology, Writing – Review & Editing. T.R.S.: Methodology, Writing – Review & Editing. F.Q.: Conceptualization, Data curation, Formal analysis, Funding acquisition, Methodology, Project administration, Resources, Supervision, Validation, Writing – Original draft, Writing – Review & Editing. B.C.S.: Conceptualization, Data curation, Formal analysis, Funding acquisition, Methodology, Project administration, Resources, Supervision, Validation, Writing – Original draft, Writing – Review & Editing.

---

**Supplementary Figure 1.**
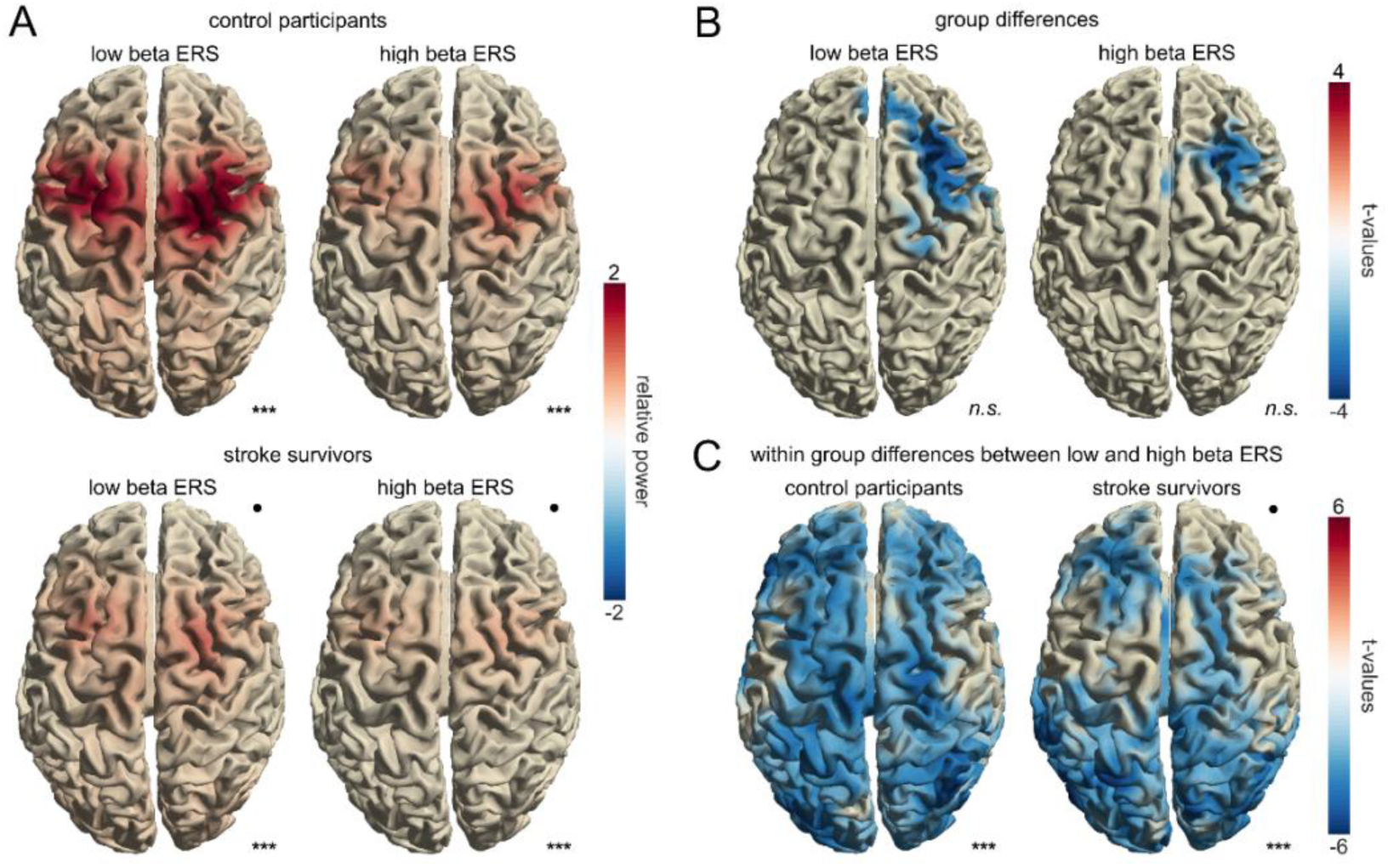
Low beta ERS power is stronger than high beta ERS power in both groups. (**A**) Beta ERS in the low beta band (14 – 20 Hz) and high beta band (21 – 29 Hz) for each group. Permutation-based cluster statistics revealed significant low as well as high beta ERS in both groups compared to baseline (control participants: low beta ERS: *p* < 0.001, high beta ERS: *p* < 0.001, stroke survivors: low beta ERS: *p* < 0.0001, high beta ERS: *p* < 0.0001, not corrected for multiple comparisons). (**B**) Results of the permutation-based cluster statistics comparing low and high beta ERS between groups. Non-significant clusters indicated a trend toward reduced low and high beta ERS in stroke survivors compared to control participants (low beta ERS: *p* = 0.13, high beta ERS: *p* = 0.15, not corrected for multiple comparisons). (**C**) Low beta ERS power was significantly stronger than high beta ERS power in both groups, as revealed by permutation-based cluster statistics (control participants: *p_cor_* < 0.001, stroke survivors: *p_cor_* < 0.001, corrected for two comparisons). The black dot represents the lesion side (right hemisphere). Significance markers: *n.s.* = not significant, \*\*\**p* < 0.001.

**Supplementary Figure 2.**
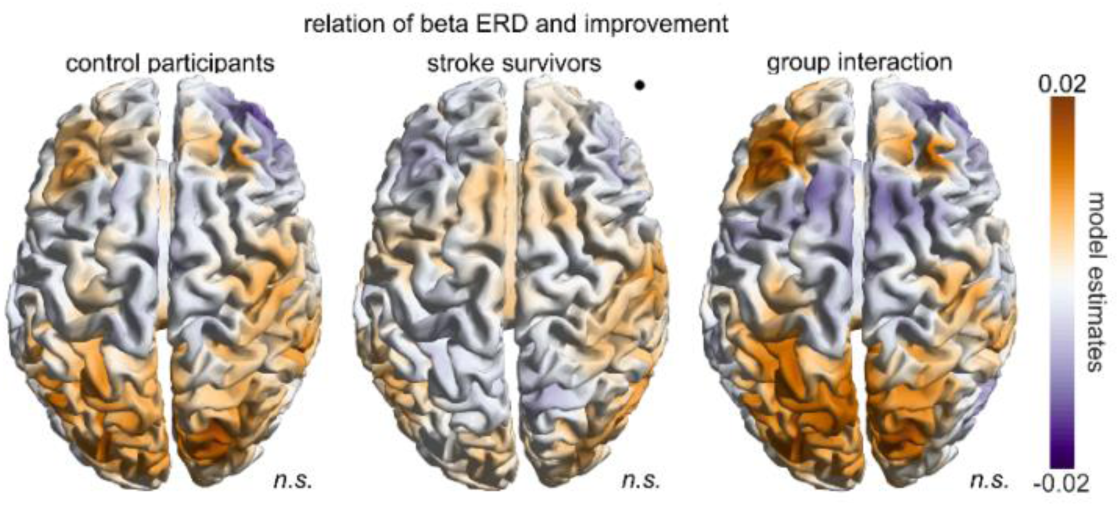
Beta ERD was not significantly linked to *improvement*. Model estimates for the factor *improvement* (model: *beta ERD ∼ performed hand + movement rate block 1 + improvement)*, calculated with permutation-based cluster statistic for each group, as well as the model estimates for the *improvement* x group interaction (model: *beta ERD ∼ performed hand + movement rate block 1 + improvement x group)*. No significant clusters were found (control participants: *p* = 0.44, stroke survivors: *p* = 0.39, interaction model: no cluster found, not corrected for multiple comparisons). The black dot represents the lesion side (right hemisphere). Significance markers: *n.s.* = not significant.

**Supplementary Figure 3.**
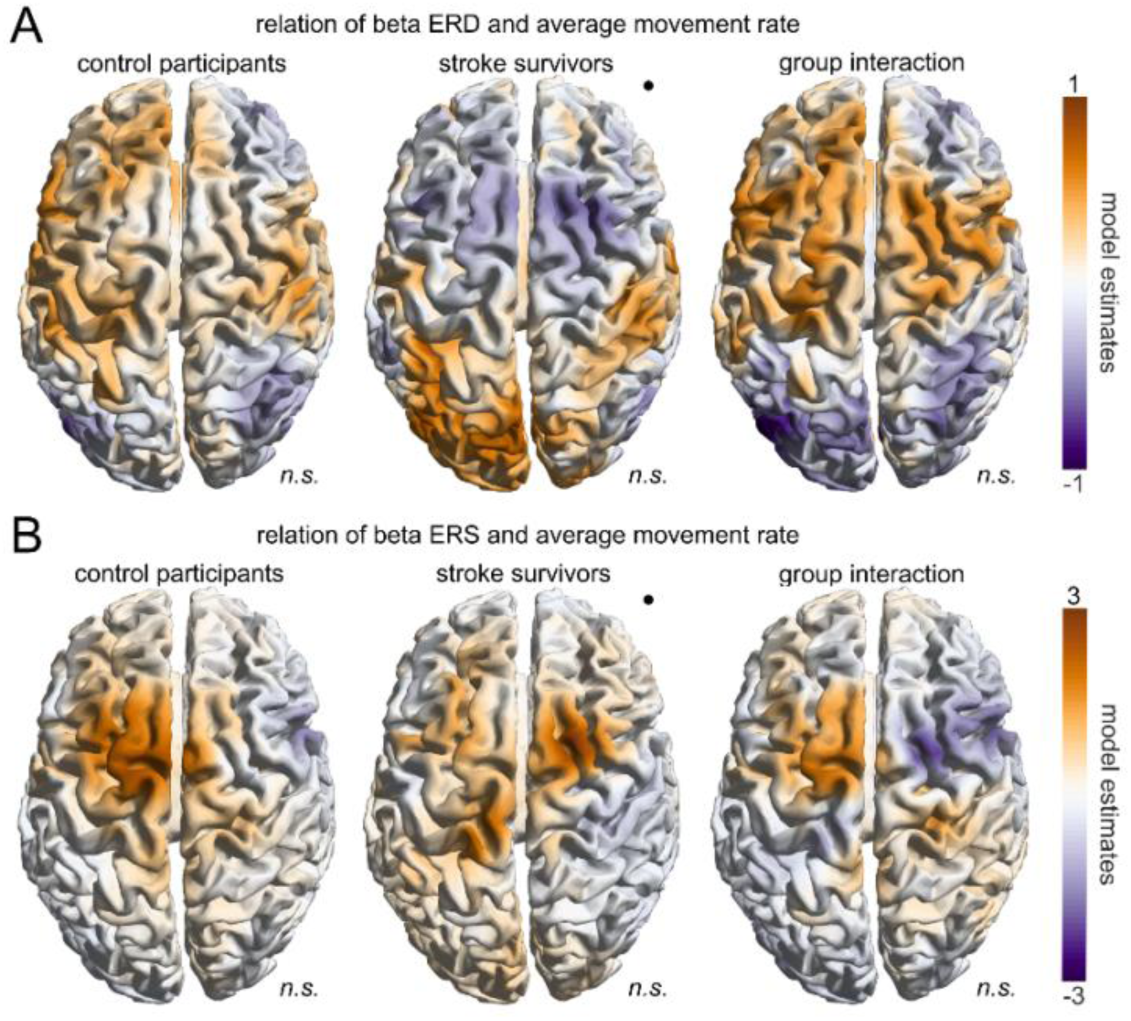
Beta ERD and beta ERS were not significantly related to *movement rate*. (**A**) Model estimates for factor average *movement rate* (model: *beta ERD ∼ performed hand + average movement rate)*, calculated with permutation-based cluster statistic for each group, as well as the model estimates of the interaction *movement rate* x group (model *beta ERD ∼ performed hand + average movement rate x group)*. No significant clusters were found (control participants: *p* = 0.44, stroke survivors: *p* = 0.29, interaction model: 0.46, not corrected for multiple comparisons). (B) Model estimates for the factor *movement rate* (model: *beta ERS ∼ performed hand + average movement rate)* calculated with permutation-based cluster statistic for each group, as well as the model estimates of the interaction *movement rate* x group (model *beta ERS ∼ performed hand + average movement rate x group)*. No significant clusters were found (control participants: *p* = 0.34, stroke survivors: *p* = 0.15, interaction model: 0.43, not corrected for multiple comparisons). The black dot represents the lesion side (right hemisphere). Significance markers: *n.s.* = not significant.

**Supplementary Table 1.**
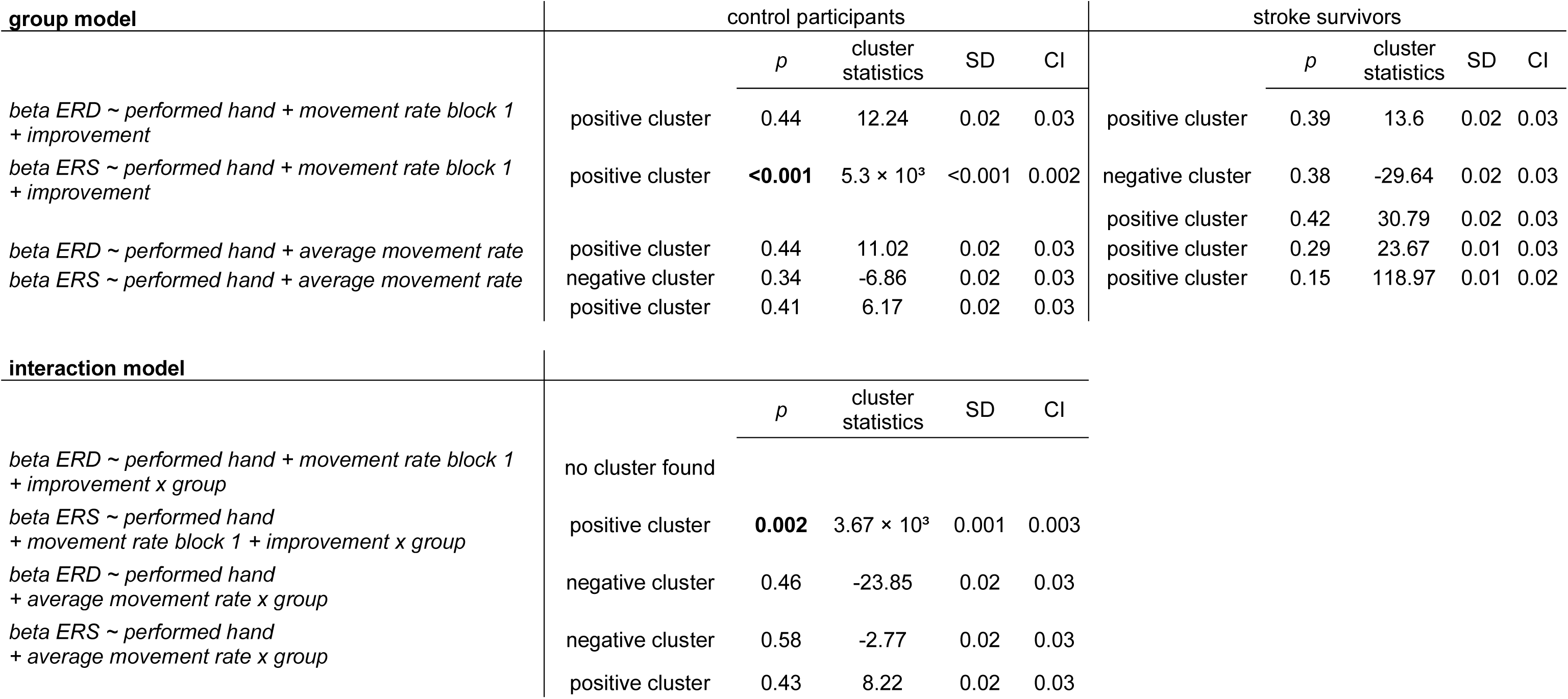
Results of linear-mixed effects model calculated with cluster-based permutation statistics on the whole brain MEG data. Not corrected for multiple comparisons.

**Supplementary Table 2.** Exploratory analysis reveals a significant interaction of BBT with group and improvement. The model “cluster ERS ∼ performed hand + movement rate block 1 + improvement x group x parameter” was calculated for each parameter (FMUE, ARAT, BBT, NHPT, grip force, key grip force, CST FA ratio, age). The standard deviations (SE) and p-values (p) are displayed for each predictor variable. Additionally, R-squared, F-statistics and p-values of each model are stated. All values are uncorrected for multiple comparisons.

| Parameter | FMUE |  | ARAT |  | BBT |  | NHPT |  | Grip force |  | Key grip |  | CST FA ratio |  | Age |  |
| --- | --- | --- | --- | --- | --- | --- | --- | --- | --- | --- | --- | --- | --- | --- | --- | --- |
|  | SE | p | SE | p | SE | p | SE | p | SE | p | SE | p | SE | p | SE | p |
| Intercept | 1.5E+02 | 6.4E-02 | 2.4E+01 | 9.8E-01 | 5.0E+00 | **4.9E-03** | 4.1E+00 | 1.9E-01 | 1.5E+00 | 2.7E-01 | 4.2E+00 | 1.4E-01 | 4.6E+00 | 4.6E-01 | 3.0E+00 | 9.4E-01 |
| Performed hand r | 2.8E-01 | **1.2E-04** | 3.0E-01 | **6.3E-04** | 2.8E-01 | **7.1E-04** | 2.9E-01 | **3.5E-02** | 3.4E-01 | **2.7E-03** | 2.8E-01 | **1.5E-03** | 4.0E-01 | **1.1E-02** | 2.6E-01 | **1.1E-03** |
| Performance block 1 | 1.1E+00 | **3.3E-02** | 1.0E+00 | 3.3E-01 | 8.7E-01 | 4.7E-01 | 8.7E-01 | 9.9E-01 | 1.1E+00 | 3.8E-01 | 1.0E+00 | 3.0E-01 | 1.0E+00 | 2.6E-01 | 8.8E-01 | 3.3E-01 |
| Improvement x Group<br>stroke x Parameter | 2.0E-01 | 7.81E-02 | NA | NA | 4.7E-03 | **1.6E-02** | 4.0E-01 | 7.1E-01 | 2.2E-01 | 5.5E-01 | 4.2E-01 | 1.4E-01 | 6.9E-01 | 7.2E-01 | 3.8E-03 | 2.6E-01 |
| R squared | 7.6E-01 |  | 7.0E-01 |  | 8.0E-01 |  | 8.2E-01 |  | 7.1E-01 |  | 7.3E-01 |  | 7.4E-01 |  | 7.9E-01 |  |
| F statistics | 6.7E+00 |  | 5.9E+00 |  | 8.6E+00 |  | 9.3E+00 |  | 5.2E+00 |  | 5.8E+00 |  | 4.8E+00 |  | 7.8E+00 |  |
| Model p-value | **2.5E-04** |  | **6.1E-04** |  | **4.7E-05** |  | **2.6E-05** |  | **1.2E-03** |  | **6.1E-04** |  | **3.7E-03** |  | **9.1E-05** |  |

